# Estimating area-level variation in SARS-CoV-2 infection fatality ratios

**DOI:** 10.1101/2021.12.04.21267288

**Authors:** Joshua Ladau, Chaincy Kuo, Eoin L. Brodie, Nicola Falco, Ishan Bansal, Elijah B. Hoffman, Marcin P. Joachimiak, Ana M. Mora, Angelica M. Walker, Haruko M. Wainwright, Yulun Wu, Daniel Jacobson, Matthias Hess, James B. Brown, Katrina Abuabara

## Abstract

**Background:** During a pandemic, estimates of geographic variability in disease burden are important but limited by the availability and quality of data.

**Methods:** We propose a framework for estimating geographic variability in testing effort, total number of infections, and infection fatality ratio (IFR). Because symptomatic people are more likely to seek testing, we use a noncentral hypergeometric model that accounts for differential probability of positive tests. We apply this framework to the United States (U.S.) COVID-19 pandemic to estimate county-level SARS-CoV-2 IFRs from March 1, 2020 to October 31, 2020. Using data on population size, number of observed cases, number of reported deaths in each U.S. county and state, and number of tests in each U.S. state, we develop a series of estimators to identify the number of SARS-CoV-2 infections and IFRs at the county level. We then perform a simulation and compare the estimated values to simulated values to demonstrate the validity of our approach.

**Findings:** Applying the county-level estimators to the real, unsimulated COVID-19 data spanning March 1, 2020 to October 31, 2020 from across the U.S., we found that IFRs varied from 0 to 0.0273, with an interquartile range of 0.0022 and a median of 0.0018. The estimators for IFRs, number of infections, and number of tests showed high accuracy and precision; for instance, when applied to simulated validation data sets, across counties, Pearson correlation coefficients between estimator means and true values were 0.88, 0.95, and 0.74, respectively.

**Interpretation:** We propose an estimation framework that can be used to identify area-level variation in IFRs and performs well to estimate county-level IFRs in the U.S. COVID-19 pandemic.

## 1 Introduction

During the course of an epidemic, accurate estimation of the infection fatality ratio (“IFR”) – defined here as the number of deaths caused by an infection per 1000 infected people – is crucial for policy and planning purposes. IFRs may vary by age distribution of the population, distribution of infection across demographic groups, prevalence of underlying health conditions in the population, access to healthcare resources, and other factors^1,2^. Therefore, a flexible framework for estimating IFRs in different geographic regions and among different populations is important.

Accurately ascertaining both the number of deaths and the number of infections can be challenging^3^. While the number of deaths may be under- or over-reported in some settings^4^, even more concerning are potential inaccuracies in the ascertainment of the number of infections. While case mortality data may provide an upper bound for the IFR, the total number of infections is likely to be underestimated because infected people may not be tested or report illness. In addition, the availability and uptake of testing, as well as reliability of testing data, may also vary over time and geography. For example, during the COVID-19 pandemic, the number of SARS-CoV-2 nucleic acid amplification tests (NAATs) administered was consistently recorded at the state level across the U.S., though not at the county level^5^. Focusing on the COVID-19 pandemic in the U.S., here we develop an inferential framework for estimating area-level variation in testing efforts, number of SARS-CoV-2 infections, and SARS-CoV-2 IFRs. Although this framework was developed to address the need for county-level SARS-CoV-2 IFR estimates in the U.S., it can readily be applied to other geographic settings and pandemics to estimate area-level variation in outcomes.

## 2 Methods

This analysis aims to develop estimators for the SARS-CoV-2 IFRs – defined here as the number of COVID-19 deaths per 1000 SARS-CoV-2 infections – within each U.S. county. Here we focus on data from March 1, 2020 - October 31, 2020, though this approach can be applied to any time period within the pandemic and to diseases other than COVID-19. There are two primary challenges with developing these estimators for COVID-19. First, the total number of SARS-CoV-2 infections is unknown because most asymptomatic individuals, but also many symptomatic individuals, fail to get tested or report illness^6,7,8^. Second, although reliable data are available on the number of SARS-CoV-2 NAATs administered per state, reliable testing data are not available at the county level^5,9^. Having data on testing is needed because it allows sampling effort to be factored in; without it, for instance, it is impossible to know whether observing one or a few cases in a county is indicative of a low SARS-CoV-2 infection ratio or just a limited testing effort. What follows is an inferential framework for estimating the testing effort, number of SARS-CoV-2 infections, and ultimately SARS-CoV-2 IFRs for each county.

### 2.1 Definitions

At a given time and for a given region (county, state, or the U.S.), let *D* denote the number of COVID-19 deaths, *C* denote the number of observed COVID-19 cases, and *T* denote the number of SARS-CoV-2 NAATs administered. Let *P* be the population size of the region, assumed effectively fixed over the timescale of the pandemic. Let the number of infections (both observed and unobserved) be denoted *ι*. Let the IFR, defined as the number of COVID-19 deaths per 1000 SARS-CoV-2 infections, be denoted *ϕ*. Define *ω* as the odds ratio of testing an infected vs an uninfected individual. County-, state-, and country-level (nation-level) variables will be denoted with the subscripts *C, S*, and *N* respectively. For instance the number of observed cases in a state will be denoted *C*_*s*_. Estimators will be denoted with ‘hats;’ e.g., the estimator for the number of infections in a county will be denoted 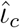. Where it is necessary to distinguish an individual county or state, an additional subscript *j* will be used; e.g., *T*_0*j*_ denotes the number of SARS-CoV-2 NAATs administered in county *j*.

### 2.2 Assumptions

To derive an estimator for the county-level IFR, assumptions about several quantities are necessary:

1. Odds ratios (*ω*): Although it is not necessary to assume actual values of *ω*, it is necessary to assume a geographic structure for them for each time point. Here we assume that at a given time point in the pandemic, the odds ratio of testing an infected vs an uninfected individual is the same for all counties, states, and the entire U.S. That is, *ω*_*c*_ = *ω*_*s*_ = *ω*_*n*_. However, the odds ratios can change through time.
2. Global IFR (*ϕ*_*n*_): To estimate geographic variation in IFR, it is necessary to assume an baseline IFR for the entire region under consideration. In our case, we assume that the IFR across the entire U.S. was five deaths per 1000 infections from March 1, 2020 to October 31, 2020, a widely agreed upon value^10^.
3. Observed quantities: we assume that the population (*P*) and numbers of COVID-19 deaths (*D*) and cases (*C*) in each county, state, and across the entire U.S. are known. Likewise, we assume that the number of tests in each state (*T*_*s*_) and consequently across the whole U.S. (*T*_*n*_) is known. However, we assume that the number of tests in each county is unknown (*T*_*c*_). While the latter assumption may seem peculiar in light of the assumption that *T*_*s*_ is known, it reflects the situation that, in the U.S., the number of SARS-CoV-2 NAATs in each county was not consistently recorded during much of 2020, despite the fact that it was recorded in aggregate for each state.
4. Number of tests in each county and state (*T*_*c*_ and *T*_*s*_): Because the number of NAATs in each county is unknown, it is necessary to estimate it. To do so, we assume that the number of tests administered *T*_*cj*_ in county *j* is related to the population size *P*_*cj*_ and the number of observed cases *C*_*cj*_ by 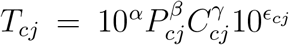, for some constants *α, β*, and *γ*, where *ϵ*_*cj*_ is distributed normally with variance zero. Thus, log *T*_*cj*_ is a linear function of *P*_*cj*_ and *C*_*cj*_: log*T*_*cj*_ = *α* + *β* log*P*_*cj*_ + *γ* log *C*_*cj*_ + *ϵ*_*cj*_. We make an analogous assumption for states, i.e., that 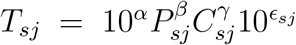, implying that log *T*_*sj*_ = *α* + *β* log *P*_*sj*_ + *γ* log *C*_*sj*_ + *ϵ*_*sj*_. Empirically, these assumptions are strongly supported for states (Figure 1). Furthermore, it reasonable to suppose that for some non-negative constants *κ* and *η*, which are additionally upper-bounded by 1: *T*_*ij*_ = *κP*_*ij*_ and *C*_*ij*_ = *ηT*_*ij*_, which is to say, that (a) the number of tests in state or county *j* is directly proportional to the population size of that state or county and (b) the number of observed cases in a state or county is directly proportional to the number of tests conducted there. Multiplying these two equations and simplifying yields:

**Figure 1:**
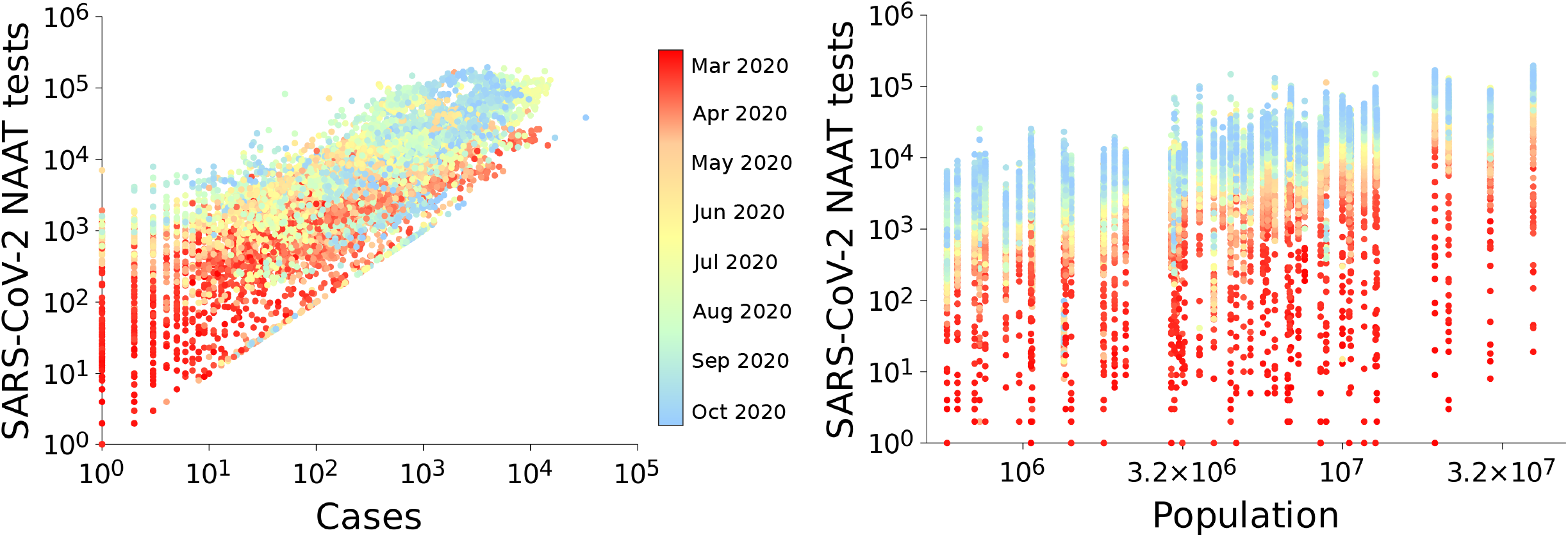
Relationship between the number of NAATs performed and the total number of COVID-19 cases and population size at the state level. Although the intercepts and slopes of these relationships varied over time, all are approximately linear when log-transformed. By applying linear models, fitted using state-level data, for the number of log tests at each time point as function of the log cases and log population size, we inferred the number of tests in each county at each time point. This inference was possible because, although in most counties the number of tests was not recorded, the number of COVID-19 cases and population size were available for all counties.

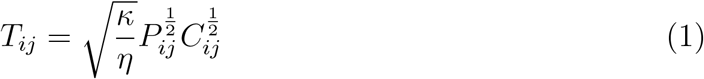

That is, the power function assumed above.

At a given time point within a specific region, it is reasonable to conceptualize the process by which people are tested with a noncentral hypergeometric model, wherein each test is administered to either a person having or not having COVID-19 with some bias toward administering tests to symptomatic people (e.g., because sick people are more likely to seek testing^8^; quantified by *ω*). For estimation, we utilize an approximation for the mean of the Wallenius’ noncentral hypergeometric distribution (Equation 16 in reference^11^):

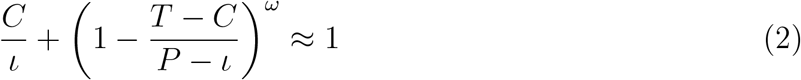

While recognizing that this expression is a close approximation, for clarity, in the following we will refer to it as an equality.

### 2.3 Estimators

We develop several moments estimators based on this equation. As per Assumptions 2 and 3, whether the quantities *T* and *ϕ* are observed and unobserved, and consequently need to be estimated, differs between regions (summarized in Table 1). We first find estimators for *ι* and *ω* at the country level, and then use these estimators to find estimators at the state and county levels (Table 3). Specifically, the estimators are derived in the following sequence:

**Table 1:**
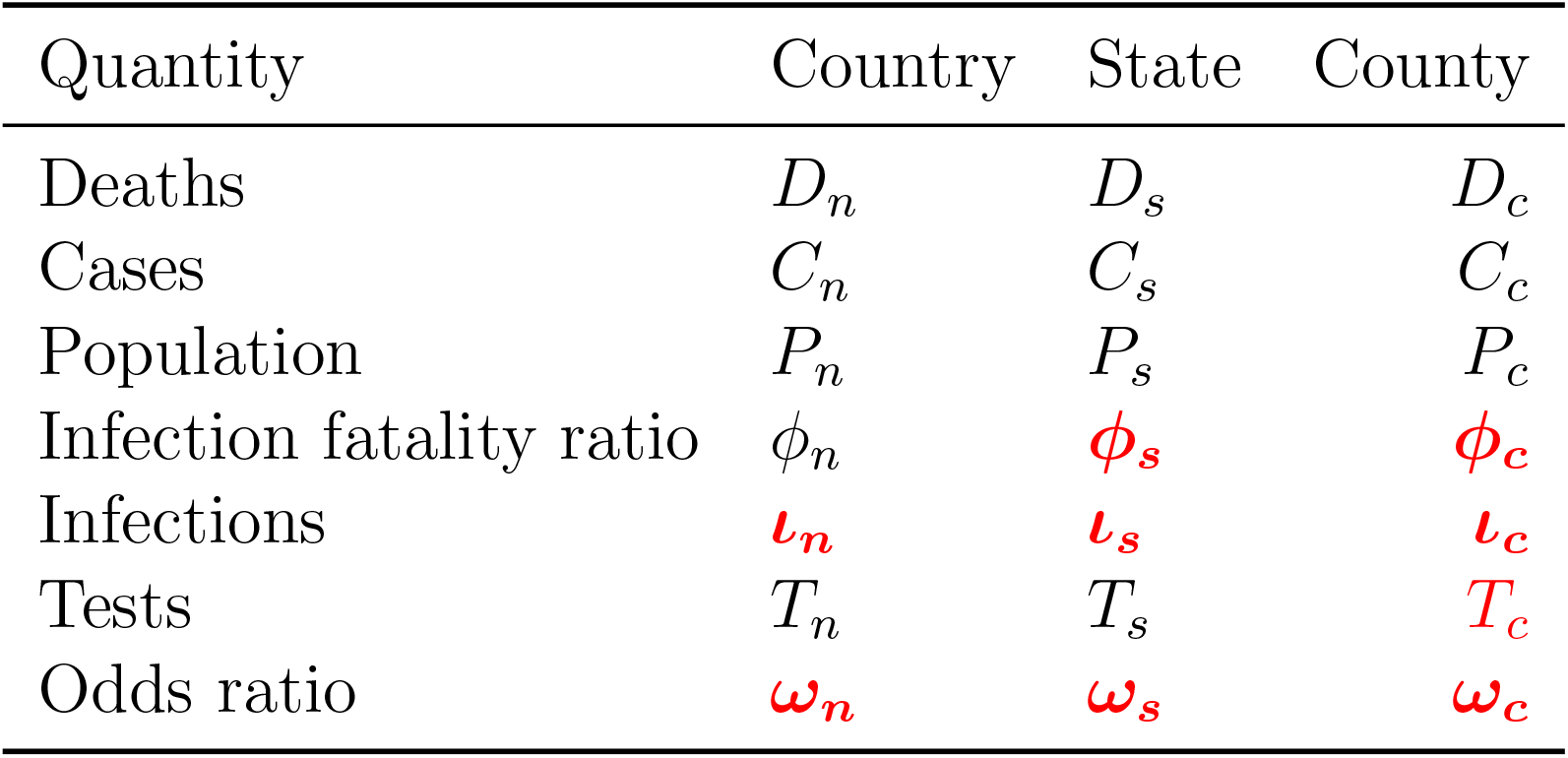
Real world COVID-19 data: Observed (black) and unobserved (red) quantities.

| Quantity | Country | State | County |
| --- | --- | --- | --- |
| Deaths | $D_n$ | $D_s$ | $D_c$ |
| Cases | $C_n$ | $C_s$ | $C_c$ |
| Population | $P_n$ | $P_s$ | $P_c$ |
| Infection fatality ratio | $\phi_n$ | $\phi_s$ | $\phi_c$ |
| Infections | $\iota_n$ | $\iota_s$ | $\iota_c$ |
| Tests | $T_n$ | $T_s$ | $T_c$ |
| Odds ratio | $\omega_n$ | $\omega_s$ | $\omega_c$ |

1. Estimate the total number of SARS-CoV-2 infections in the country using Assumption 2 (i.e., that the IFR is five deaths per 1000) and the fact that the number of deaths is known.
2. Estimate the odds ratio at the county level by substituting the number of infections estimated in 1 and solving Equation (2) for *ω*.
3. Estimate the odds ratio at the state level via the Assumption 1 listed above.
4. Estimate the number of SARS-CoV-2 infections at the state level using the odds ratio from the previous step and solving Equation (2) for *ι*.
5. Estimate the IFR at the state level by dividing the number of deaths (observed) at the state level by the number of SARS-CoV-2 infections inferred in the previous step.
6. Estimate the odds ratio at the county level via Assumption 1 listed above.
7. Estimate the number of tests performed in each county using Assumption 4 listed above.
8. Estimate the number of SARS-CoV-2 infections at the county level using the odds ratio and tests estimates from the previous two steps, and solving Equation (2) for *ι*.
9. Estimate the IFR at the county level by dividing the number of COVID-19 deaths (observed) at the county level by the number of SARS-CoV-2 infections inferred in the previous step.

### 2.4 Validation and application

We simulated COVID-19 case data from March 1, 2020 to October 31, 2020, and then numerically assessed the performance of the estimators. Specifically, for each week of the study period:

1. We set the country-level odds ratio, *ω*_*n*_ equal to 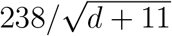, where *d* is the number of days elapsed since March 1, 2020. (With regard to the numerator, there were 238 days between March 1, 2020 and October 31, 2020.) As per Assumption 1, we set the state- and county-level odds ratios equal to the same country-level values.
2. For each region, with the exception of the numbers of cases and country-wide IFR, we set the observed quantities in Table 1 equal to their known values (Table 2).
3. We simulated the numbers of tests in counties, *T*_*c*_, by distributing the tests in the corresponding states (*T*_*s*_; an observed quantity) according to a multivariate hypergeometric distribution, with the condition that the number of simulated tests in a county could not exceed its population size (*P*_*c*_; also an observed quantity).
4. For each county, we simulated the total number of SARS-CoV-2 infections (*ι*_*c*_) as a random variate from the following distribution: ⌊0.017 *· P*_*c*_*U* ^4^⌋, where ⌊*·* ⌋ is the floor function and *U* is a uniform [0, 1] random variate, with the condition that *ι*_*c*_ ≥*D*_*c*_. While this may seem like an arbitrary choice of distributions, it has the desirable property of resulting in a country-wide IFR (*ϕ*_*n*_) of approximately 5 deaths per 1000 infections, consistent with Assumption 2.
5. For each county, we simulated the number of COVID-19 cases in each county (*C*_*c*_) as a variate from Wallenius’ noncentral hypergeometric distribution, using the simulated odds ratio and number of SARS-CoV-2 infections from above.
6. For each state and the entire country, we found simulated numbers of SARS-CoV-2 infections and COVID-19 cases (*ι*_*s*_, *ι*_*n*_, *C*_*s*_, and *C*_*n*_) by summing the corresponding values simulated above from the counties that it contained.

Using the simulated values of the known quantities in the original data set (black quantities in Table 1) and the estimators described above, we found estimates of all of the unknown quantities (red quantities in Table 1), and also the country-wide IFR, *ϕ*_*n*_. The average and variance of the estimates over 100 iterations of the simulation process yielded measures of the performance of the estimators.

## 3 Results

Applying the county-level estimators to the real, unsimulated COVID-19 data spanning March 1, 2020 to October 31, 2020 from across the U.S., we found that IFRs varied from 0 to 0.0273, with a mean of 0.0023 and a median of 0.0018 (interquartile range 0.0022). IFRs were the lowest in the Intermountain West, High Plains, and New England, but the highest in Arizona and Western New Mexico, the Upper Midwest, and scattered other regions of the U.S. The number of SARS-CoV-2 infections (not corrected for population) was highest in the Southwest and scattered other parts of the U.S., and lowest in Utah. Testing (also not corrected for population) was generally highest around large population centers. Similar trends were evident at the state level, with lower spatial resolution (Figure 2).

**Table 2:** Validation data: Observed (black) and simulated (blue) quantities. Starred quantities were simulated as being non-random.

| Quantity | Country | State | County |
| --- | --- | --- | --- |
| Deaths | $D_n$ | $D_s$ | $D_c$ |
| Cases | $\textcolor{blue}{C}_n$ | $\textcolor{blue}{C}_s$ | $\textcolor{blue}{C}_c$ |
| Population | $P_n$ | $P_s$ | $P_c$ |
| Infection fatality ratio | $\textcolor{blue}{\phi}_n$ | $\textcolor{blue}{\phi}_s$ | $\textcolor{blue}{\phi}_c$ |
| Infections | $\textcolor{blue}{\iota}_n$ | $\textcolor{blue}{\iota}_s$ | $\textcolor{blue}{\iota}_c$ |
| Tests | $T_n$ | $T_s$ | $T_c^*$ |
| Odds ratio | $\textcolor{blue}{\omega}_n^*$ | $\textcolor{blue}{\omega}_s^*$ | $\textcolor{blue}{\omega}_c^*$ |

**Table 3:** Estimators used for finding the unobserved quantities listed in Table 1. Bold numbers indicate the order of calculation. The variables 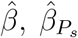, and 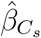 in **(7)** are linear regression coefficient estimates resulting from regressing log *T*_*s*_ on log *P*_*s*_ and log *C*_*s*_. The estimators for the total number of infections at the state and county levels [**(4)** and **(8)**; 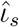 and 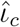] are defined implicitly and are found by solving the given equations numerically. These estimators and 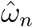 are based on the approximation for the mean of the Wallenius’ noncentral hypergeometric distribution given in Equation (2). Time (*t*), state (*j*), and county (*i*) subscripts are omitted for readability, but all estimators are time-, state-, and county-specific.

| Region | Infection fatality ratio | Infections | Tests | Odds ratio |
| --- | --- | --- | --- | --- |
| Country | $\phi_n = 5$ (observed) | **(1)** $\hat{\iota}_n = \frac{1000D_n}{\phi_n}$ | $T_n$ (observed) | **(2)** $\hat{\omega}_n = \frac{\log(1 - \frac{C_n}{\hat{\iota}_n})}{\log(1 - \frac{T_n - C_n}{P_n - \hat{\iota}_n})}$ |
| State | **(5)** $\hat{\phi}_s = \frac{1000D_s}{\hat{\iota}_s}$ | **(4)** $\frac{C_s}{\hat{\iota}_s} + \left(1 - \frac{T_s - C_s}{P_s - \hat{\iota}_s}\right)^{\hat{\omega}_s} = 1$ | $T_s$ (observed) | **(3)** $\hat{\omega}_s = \hat{\omega}_n$ |
| County | **(9)** $\hat{\phi}_c = \frac{1000D_c}{\hat{\iota}_c}$ | **(8)** $\frac{C_c}{\hat{\iota}_c} + \left(1 - \frac{\hat{T}_c - C_c}{P_c - \hat{\iota}_c}\right)^{\hat{\omega}_c} = 1$ | **(7)** $\hat{T}_c = e^{\hat{\beta} + \hat{\beta}_{P_s} \log P_c + \hat{\beta}_{C_s} \log C_c}$ | **(6)** $\hat{\omega}_c = \hat{\omega}_n$ |

**Figure 2:**
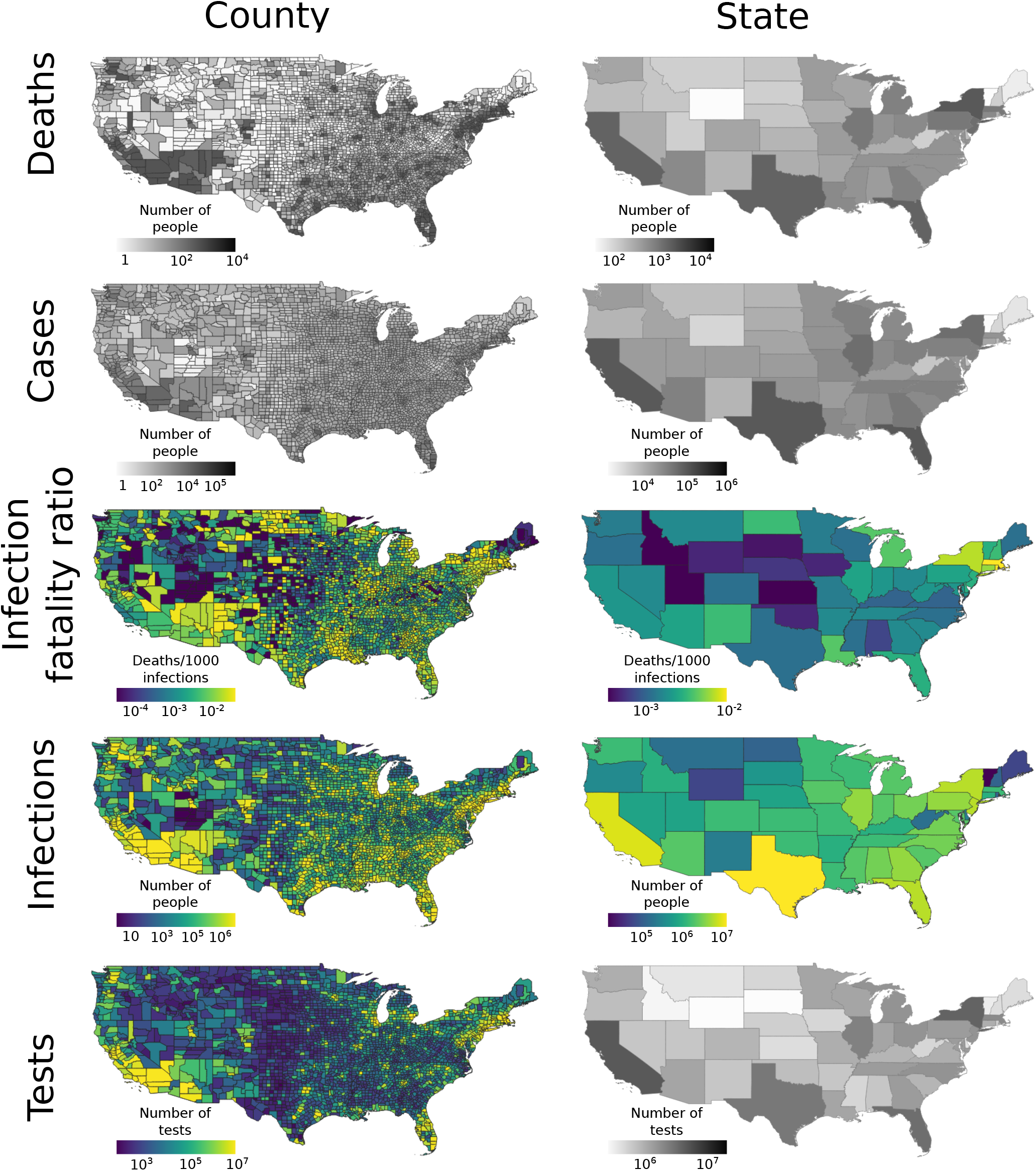
Maps showing estimated and observed values of the number of deaths, cases, infection fatality ratios, number of infections, and number of tests in the U.S. over the period spanning March 1, 2020 to October 31, 2020. Estimated quantities are shown in color, while observed quantities are shown in gray. The number of tests was observed at the state level but unobserved at the county level, where it is estimated.

In our validation, we found that the mean of the county-level estimates of IFRs, number of SARS-CoV-2 infections, and tests had high accuracy (Figure 3; Pearson correlation coefficients for estimator means and true values 0.88, 0.95, and 0.74, respectively). Likewise, at the state level, the mean of the estimates for IFRs and number of infections had high accuracy and precision (Figure 4; Pearson correlation coefficients for estimator means and true values 0.95 and 0.99, respectively). Moreover, the estimators had coefficients of variation below 0.1 for almost all counties and states (Figure 5; for example, for IFR, 98.4%, 92.3%, and 11.6% of counties had coefficients of variation below 0.1, 0.05, and 0.01, respectively). In contrast, naive IFR estimates obtained simply by using the numbers of cases and case fatality ratio were biased by up to several orders of magnitude and had low precision (Figure 6; Pearson correlation coefficients for estimator means and true values were 0.21 and 0.51, respectively).

**Figure 3:**
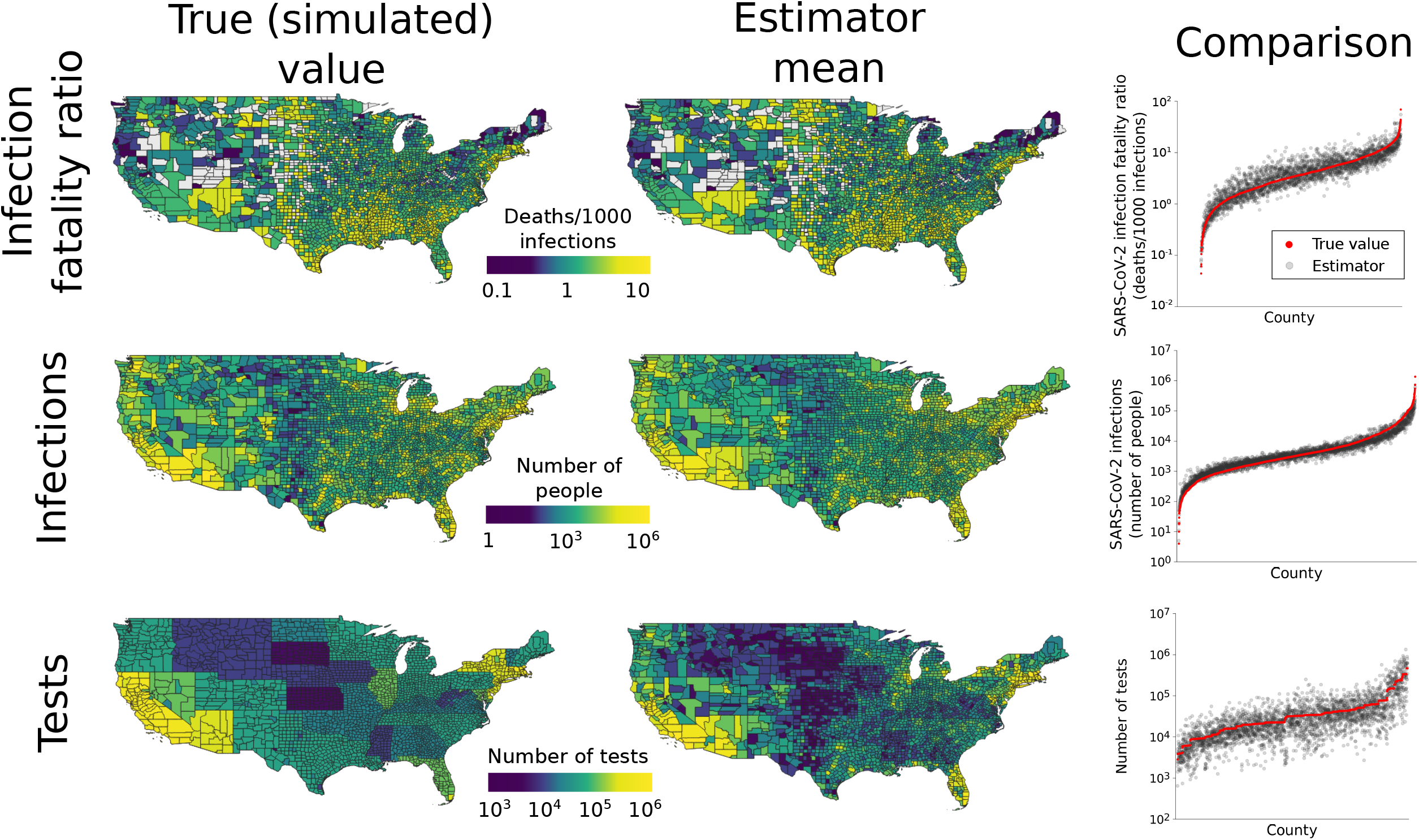
Performance of county-level estimators. Maps in the first column show true (simulated) infection fatality ratios, numbers of infections, and tests in each county. All values were conditioned on observed numbers of deaths. Counties with zero deaths are omitted from the IFR estimation results because they have ratios of zero. The second column gives the means of the estimates over 100 simulated data sets for each county. The close match between true values and estimates indicates that the estimators have high accuracy. The accuracy is further illustrated by the graphs in the third column, which plot the true values in each county compared to the mean of the estimator. In each plot, counties (*x*-axis) are ordered by increasing value.

**Figure 4:**
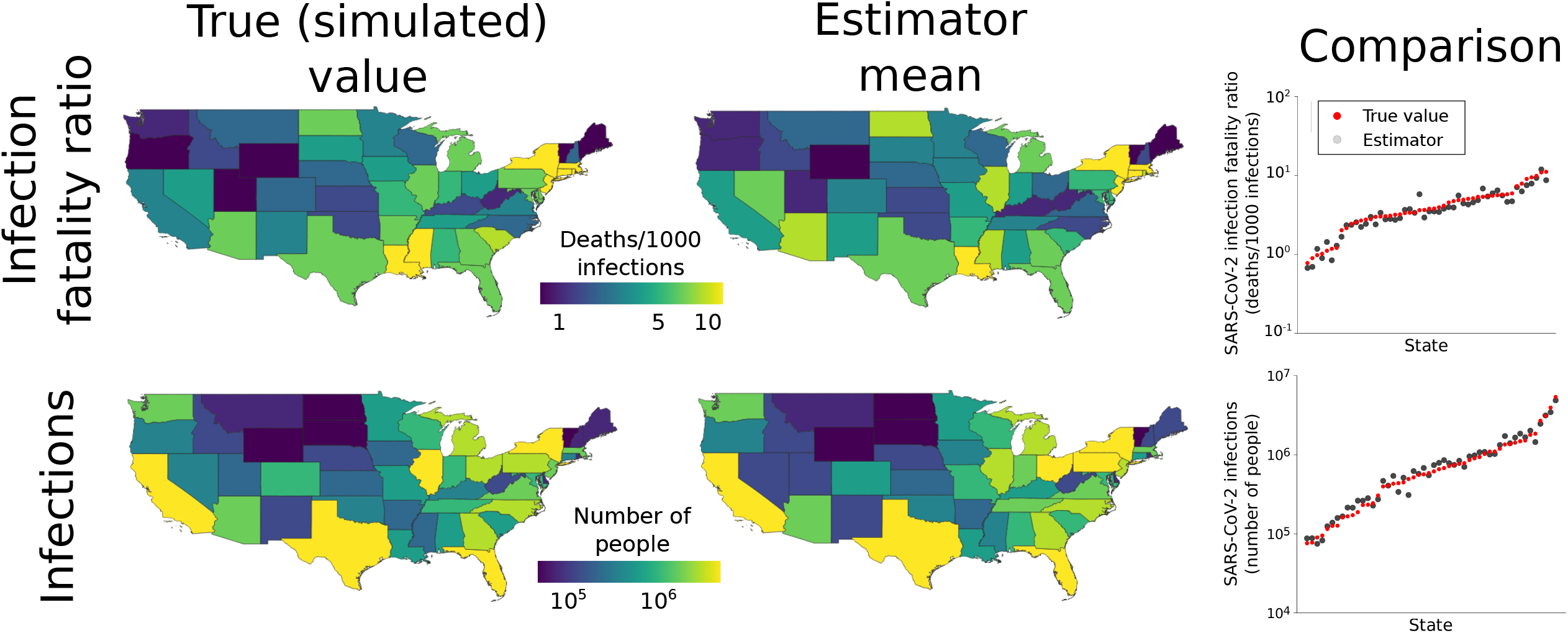
Performance of state-level estimators. The maps and graphs are analogous to those in Figure 3, but are at the state level. Unlike at the county level, at the state level the the number of tests is observed rather than estimated.

**Figure 5:**
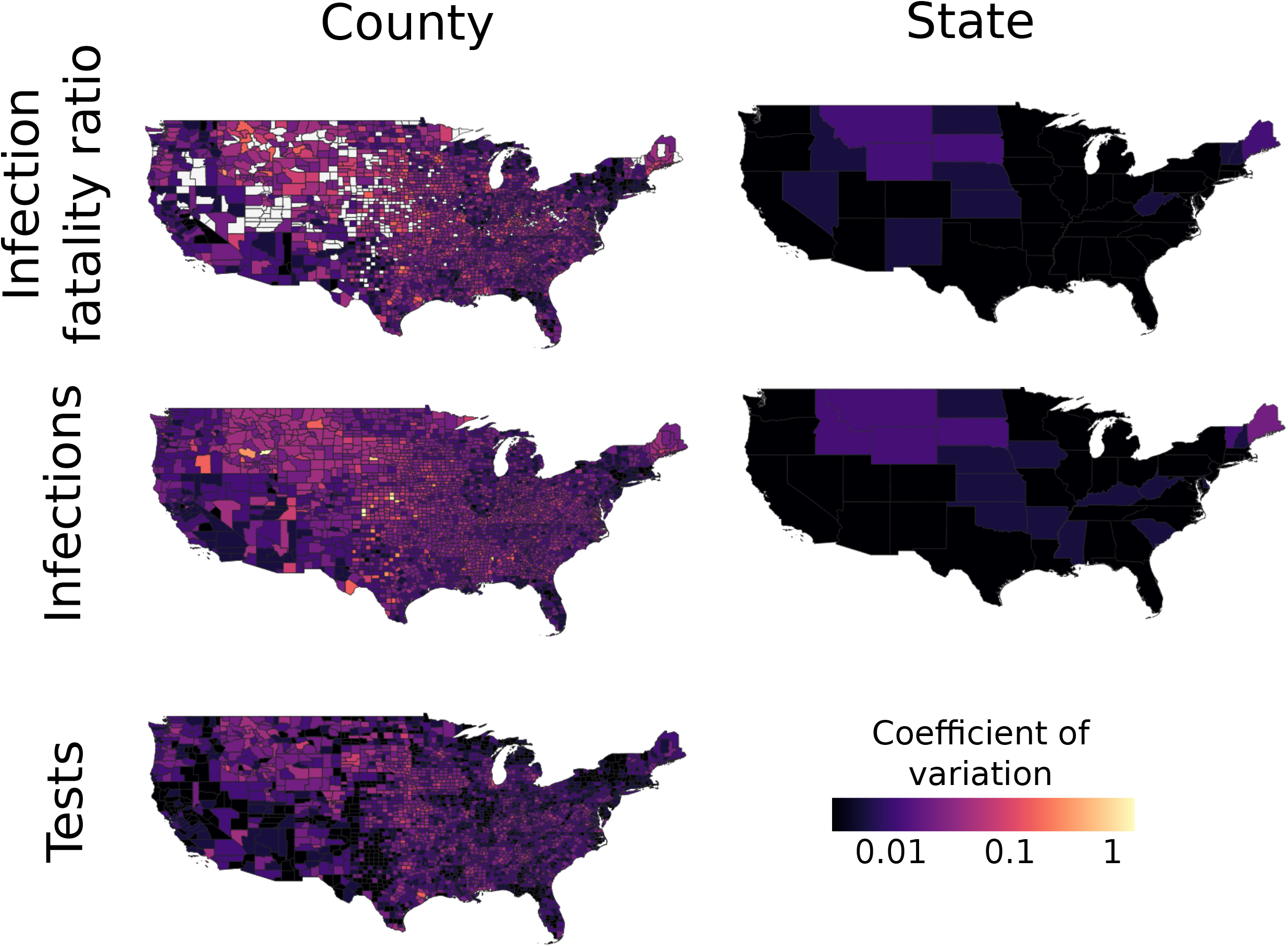
The estimators for the IFR, number of infections, and number of tests have high precision. At both the county and state levels, the estimators have low coefficients of variation, generally below 0.1 and often below 0.01.

**Figure 6:**
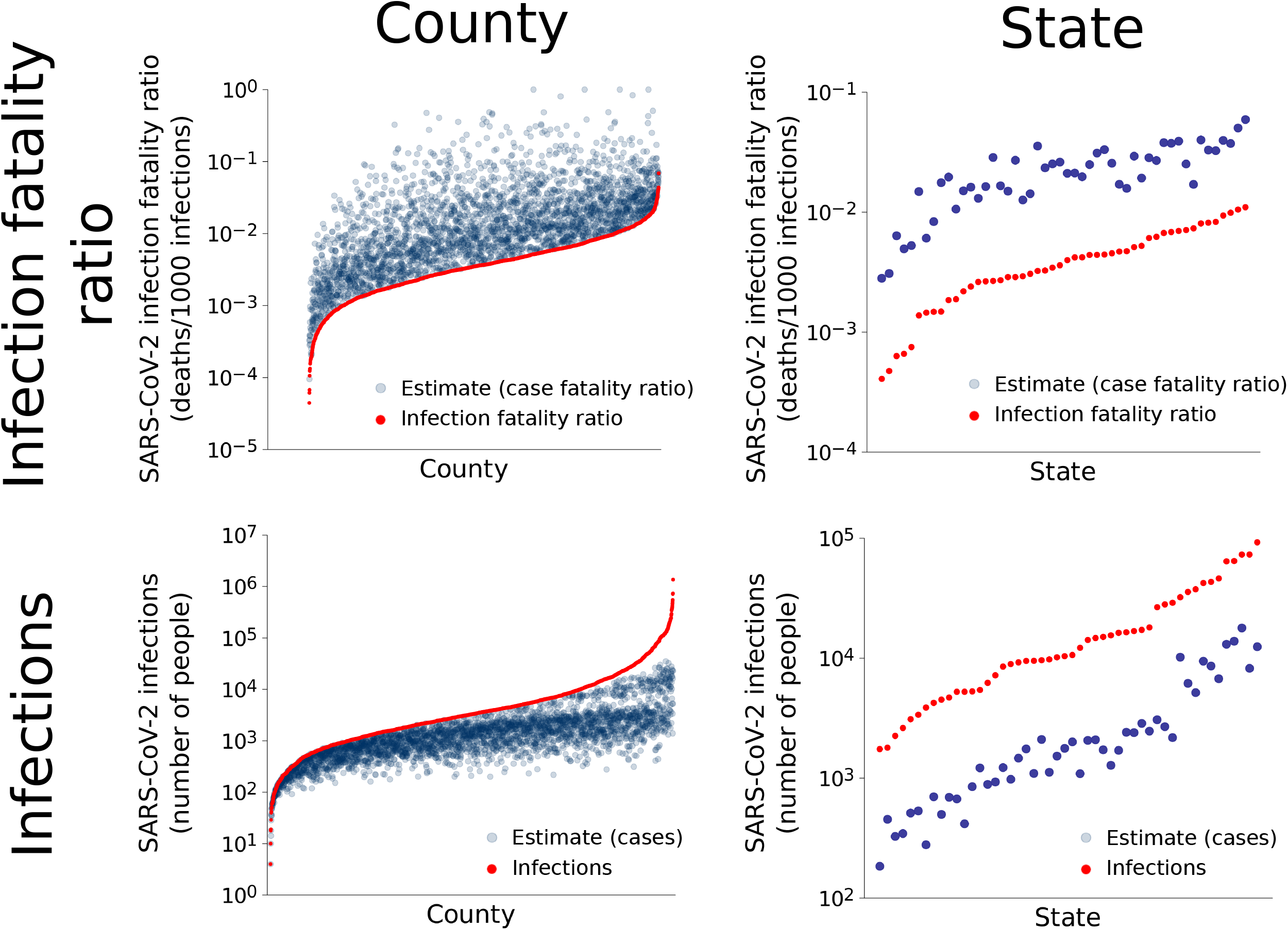
The estimators developed here greatly outperform uncorrected estimators of the IFR and number of infections, respectively. With simulated data where true values are known, at both the county and state levels, the case fatality ratio and number of cases overestimate and underestimate the IFR and number of infections, respectively, by up to several orders of magnitude. While this might seem expected, it shows that the estimators for developed here for the IFR and number of infections make substantial corrections.

## 4 Discussion

We present an inferential framework for estimating testing effort, number of SARS-CoV-2 infections, and ultimately SARS-CoV-2 IFRs in U.S. counties. The approach relies on a noncentral hypergeometric model that accounts for differential probability of positive tests (i.e. because sick individuals are more likely to seek testing than healthy individuals^8^) and performs well in validation testing.

Seroprevalence data have been widely used to estimate IFRs^12,13,14^, and in limited settings, comprehensive case reporting and tracking exist^15^. We offer the proposed framework as an extension to such methods. Our estimates require an assumption about the baseline IFR (Assumption 2), which may be based on seroprevalence or detailed tracking data. However, such data are often limited geographically and may have non-representative samples or fail to account for potential delays from onset to seroconversion^16^. Therefore, we propose using data on testing effort, where reliably reported, to address area-level and temporal variation.

The number of excess deaths is another frequently used metric of the burden of mortality potentially related to the COVID-19 pandemic. Typically defined as the difference between the observed number of deaths and the expected number of deaths across a given time period^17^, it includes changes in mortality that may directly or indirectly be attributable to COVID-19 (such as such as from delayed care or behavioral health crises). While useful in some settings, excess death estimates are highly sensitive to the reference time period used and negative estimates often occur in younger age strata^18,19^. More detailed area-level estimates of mortality directly attributable to an infection are often needed.

Similar to all models, our results are dependent on the quality of the data, which has limitations. Nonetheless, an advantage of our approach is that it allows investigators to rely on the best available data and estimate quantities where data may be lacking. Our estimates use data on NAAT (also known as molecular or PCR tests) and do not include data from antigen tests including home tests, which did not receive FDA approval until late 2020. The NAAT are the most reliable for detecting infections and were the most widely used during the study period.

Our approach relies on a number of assumptions that can be relaxed to allow for generalization. First, Assumption 1 – that the odds testing an infected vs an uninfected individual is the same for all counties, states, and the entire country – can be relaxed in numerous ways. While some geographic or temporal structure must be assumed to estimate the odds ratios, which are latent variables, this structure does not need to be geographically homogeneous. With additional covariates (e.g., data on demographics, health care access), it may also be possible to further infer specific aspects of the odds ratios. Second, regarding Assumption 2, while an overall IFR must be assumed, this value can in principle be any number between 0 and 1 dependent on the disease, time, location, and other factors. Moreover, when the aim is to rank IFR in subregions relative to each other instead of estimate their precise values, the assumed overall IFR may not matter. And third, an added degree of complexity specific to COVID-19 in the U.S. is imparted here because the number of NAATs in each state, but not county, are known. In a more typical scenario, when testing effort is recorded for each subregion, one could directly make estimates for a single class of subregions (e.g., counties) directly from a superregion (e.g., country). This would omit the need for our Assumption 4 and simplify Assumption 3. By virtue of its flexibility, our approach shows promise for application at different geographic scales and may be particularly useful in settings in which resources to carry out large, representative seroprevalence studies are not available.

Our approach can be flexibly adapted over time and space. This flexibility is important because the IFR is not biologically fixed. It varies with the distribution of age, health, and other qualities of those infected as well as the medical care they receive. Local estimates may be more accurate and useful for policy planning purposes than generalized or country-level estimates. For instance, population age structures do not address all of the heterogeneity between countries IFRs based on seroprevalence^13^. In conclusion, our approach can serve as an extension to other methods for estimation of pandemic burden to allow for greater precision around geographic and/or temporal population subgroups.

## Data Availability

All data produced in the present study are available upon reasonable request to the authors.

## 5 Acknowledgements

This research used resources of the Oak Ridge Leadership Computing Facility, which is a DOE Office of Science User Facility supported under Contract DE-AC05-00OR22725. This manuscript has been coauthored by UT-Battelle, LLC under contract no. DE-AC05-00OR22725 with the U.S. Department of Energy. The United States Government retains and the publisher, by accepting the article for publication, acknowledges that the United States Government retains a nonexclusive, paid-up, irrevocable, world-wide license to publish or reproduce the published form of this manuscript, or allow others to do so, for United States Government purposes. The Department of Energy will provide public access to these results of federally sponsored research in accordance with the DOE Public Access Plan (http://energy.gov/downloads/doe-public-access-plan, last accessed September 16, 2020). Work at Oak Ridge and Lawrence Berkeley National Laboratories was supported by the DOE Office of Science through the National Virtual Biotechnology Laboratory, a consortium of DOE national laboratories focused on response to COVID-19, with funding provided by the Coronavirus CARES Act, and was facilitated by previous breakthroughs obtained through the Laboratory Directed Research and Development Programs of the Lawrence Berkeley and Oak Ridge National Laboratories. M.P.J. was supported by a grant from the Laboratory Directed Research and Development (LDRD) Program of Lawrence Berkeley National Laboratory under U.S. Department of Energy Contract No. DE-AC02-05CH11231.

## References

[1] Chen Shen, Derrick VanGennep, Alexander F Siegenfeld, and Yaneer Bar-Yam. Unraveling the flaws of estimates of the infection fatality rate for COVID-19. Journal of Travel Medicine, 28(2), 01 2021. ISSN 1708-8305. doi: 10.1093/jtm/taaa239. URL https://doi.org/10.1093/jtm/taaa239.taaa239.

[2] Simon N. Wood, Ernst C. Wit, Matteo Fasiolo, and Peter J. Green. Covid-19 and the difficulty of inferring epidemiological parameters from clinical data. The Lancet Infectious Diseases, 21(1):27–28, Jan 2021. ISSN 1473-3099. doi: 10.1016/S1473-3099(20)30437-0. URL https://doi.org/10.1016/S1473-3099(20)30437-0.

[3] W Dana Flanders and David G Kleinbaum. Basic models for disease occurrence in epidemiology. International Journal of Epidemiology, 24(1):1–7, 02 1995. ISSN 0300-5771. doi: 10.1093/ije/24.1.1. URL https://doi.org/10.1093/ije/24.1.1.

[4] A pandemic primer on excess mortality statistics and their comparability across countries. https://ourworldindata.org/covid-excess-mortality. Accessed: 2021-09-10.

[5] The Atlantic. The covid tracking project, 2021. URL https://covidtracking.com.

[6] Prevalence of asymptomatic sars-cov-2 infection. Annals of Internal Medicine, 173(5): 362–367, 2020. doi: 10.7326/M20-3012. URL https://doi.org/10.7326/M20-3012. PMID: 32491919.

[7] Rita Rubin. First It Was Masks; Now Some Refuse Testing for SARS-CoV-2. JAMA, 324(20):2015–2016, 11 2020. ISSN 0098-7484. doi: 10.1001/jama.2020.22003. URL https://doi.org/10.1001/jama.2020.22003.

[8] Harlan Campbell, Perry de Valpine, Lauren Maxwell, Valentijn MT de Jong, Thomas Debray, Thomas Jänisch, and Paul Gustafson. Bayesian adjustment for preferential testing in estimating the covid-19 infection fatality rate: Theory and methods, 2020. URL https://arxiv.org/abs/2005.08459v2.

[9] A Jay Holmgren, Nate C Apathy, and Julia Adler-Milstein. Barriers to hospital electronic public health reporting and implications for the COVID-19 pandemic. Journal of the American Medical Informatics Association, 27(8):1306–1309, 06 2020. ISSN 1527-974X. doi: 10.1093/jamia/ocaa112. URL https://doi.org/10.1093/jamia/ocaa112.

[10] Megan O’Driscoll, Gabriel Ribeiro Dos Santos, Lin Wang, Derek A. T. Cummings, Andrew S. Azman, Juliette Paireau, Arnaud Fontanet, Simon Cauchemez, and Henrik Salje. Age-specific mortality and immunity patterns of sars-cov-2. Nature, 590 (7844):140–145, Feb 2021. ISSN 1476-4687. doi: 10.1038/s41586-020-2918-0. URL https://doi.org/10.1038/s41586-020-2918-0.

[11] Agner Fog. Calculation methods for wallenius’ noncentral hypergeometric distribution. Communications in Statistics - Simulation and Computation, 37(2):258–273, 2008. doi: 10.1080/03610910701790269. URL https://doi.org/10.1080/03610910701790269.

[12] John P. A. Ioannidis. Infection fatality rate of covid-19 inferred from seroprevalence data. Bulletin of the World Health Organization, 99(1):19–33F, Jan 2021. ISSN 1564-0604. doi: 10.2471/BLT.20.265892. URL https://pubmed.ncbi.nlm.nih.gov/33716331. 33716331[pmid].

[13] Megan O’Driscoll, Gabriel Ribeiro Dos Santos, Lin Wang, Derek A. T. Cummings, Andrew S. Azman, Juliette Paireau, Arnaud Fontanet, Simon Cauchemez, and Henrik Salje. Age-specific mortality and immunity patterns of sars-cov-2. Nature, 590 (7844):140–145, Feb 2021. ISSN 1476-4687. doi: 10.1038/s41586-020-2918-0. URL https://doi.org/10.1038/s41586-020-2918-0.

[14] Robert Verity, Lucy C. Okell, Ilaria Dorigatti, Peter Winskill, Charles Whittaker, Natsuko Imai, Gina Cuomo-Dannenburg, Hayley Thompson, Patrick G. T. Walker, Han Fu, Amy Dighe, Jamie T. Griffin, Marc Baguelin, Sangeeta Bhatia, Adhiratha Boonyasiri, Anne Cori, Zulma Cucunubá, Rich FitzJohn, Katy Gaythorpe, Will Green, Arran Hamlet, Wes Hinsley, Daniel Laydon, Gemma Nedjati-Gilani, Steven Riley, Sabine van Elsland, Erik Volz, Haowei Wang, Yuanrong Wang, Xiaoyue Xi, Christl A. Donnelly, Azra C. Ghani, and Neil M. Ferguson. Estimates of the severity of coronavirus disease 2019: a model-based analysis. The Lancet Infectious Diseases, 20 (6):669–677, Jun 2020. ISSN 1473-3099. doi: 10.1016/S1473-3099(20)30243-7. URL https://doi.org/10.1016/S1473-3099(20)30243-7.

[15] Andrew T. Levin, William P. Hanage, Nana Owusu-Boaitey, Kensington B. Cochran, Seamus P. Walsh, and Gideon Meyerowitz-Katz. Assessing the age specificity of infection fatality rates for covid-19: systematic review, meta-analysis, and public policy implications. European Journal of Epidemiology, 35(12):1123– 1138, Dec 2020. ISSN 1573-7284. doi: 10.1007/s10654-020-00698-1. URL https://doi.org/10.1007/s10654-020-00698-1.

[16] Saki Takahashi, Bryan Greenhouse, and Isabel Rodríguez-Barraquer. Are Seroprevalence Estimates for Severe Acute Respiratory Syndrome Coronavirus 2 Biased? The Journal of Infectious Diseases, 222(11):1772–1775, 08 2020. ISSN 0022-1899. doi: 10.1093/infdis/jiaa523. URL https://doi.org/10.1093/infdis/jiaa523.

[17] Centers for Disease Control and Prevention. Notes from the field: Update on excess deaths associated with the covid-19 pandemic — united states, january 26, 2020–february 27, 2021, 2021. URL https://www.cdc.gov/mmwr/volumes/70/wr/mm7015a4.htm.

[18] Steven H. Woolf, Derek A. Chapman, Roy T. Sabo, and Emily B. Zimmerman. Excess deaths from covid-19 and other causes in the us, march 1, 2020, to january 2, 2021. JAMA, 325(17):1786–1789, 05 2021. ISSN 0098-7484. doi: 10.1001/jama.2021.5199. URL https://doi.org/10.1001/jama.2021.5199.

[19] Excess deaths associated with covid-19. https://www.cdc.gov/nchs/nvss/vsrr/covid19/excessdeaths.htm. Accessed: 2021-09-02.

